# Development and validation of a harmonized memory score for multicenter Alzheimer’s disease and related dementia research

**DOI:** 10.1101/2025.03.31.25324964

**Authors:** Mark Sanderson-Cimino, Alden L. Gross, Leslie S. Gaynor, Emily W. Paolillo, Rowan Saloner, Marilyn S. Albert, fLiana G. Apostolova, Brooke Boersema, Adam L. Boxer, Bradley F. Boeve, Kaitlin B. Casaletto, Savannah R. Hallgarth, Valentina E. Diaz, Lindsay R. Clark, Pauline Maillard, Ani Eloyan, Sarah Tomaszewski Farias, Mitzi M. Gonzales, Dustin B. Hammers, Renaud La Joie, Yann Cobigo, Amy Wolf, Benjamin M. Hampstead, Dawn Mechanic-Hamilton, Bruce L. Miller, Gil D. Rabinovici, John M. Ringman, Howie J. Rosen, Sephira G. Ryman, Jillian L. Prestopnik, David P. Salmon, Glenn E. Smith, Charles DeCarli, Kumar B. Rajan, Lee-Way Jin, Jason Hinman, David K. Johnson, Danielle Harvey, Myriam Fornage, Joel H. Kramer, Adam M. Staffaroni, ALLFTD consortium, MarkVCID study, LEADS consortium, Diverse Vascular Contributions to Cognitive Impairment and Dementia (Diverse VCID) Study Investigators

## Abstract

**INTRODUCTION:** List-learning tasks are important for characterizing memory in ADRD research, but the Uniform Data Set neuropsychological battery (UDS-NB) lacks a list-learning paradigm; thus, sites administer a range of tests. We developed a harmonized memory composite that incorporates UDS memory tests and multiple list-learning tasks.

**METHODS:** Item-banking confirmatory factor analysis was applied to develop a memory composite in a diagnostically heterogenous sample (n=5943) who completed the UDS-NB and one of five list-learning tasks. Construct validity was evaluated through associations with demographics, disease severity, cognitive tasks, brain volume, and plasma phosphorylated tau (p-tau181 and p-tau217). Test-retest reliability was assessed. Analyses were replicated in a racially/ethnically diverse cohort (n=1058).

**RESULTS:** Fit indices, loadings, distributions, and test-retest reliability were adequate. Expected associations with demographics and clinical measures within development and validation cohorts supported validity.

**DISCUSSION:** This composite enables researchers to incorporate multiple list-learning tasks with other UDS measures to create a single metric.

## 1. Background

Alzheimer’s Disease (AD) and AD-Related Dementia (ADRD) research has been accelerated by the standardization of data collection procedures across multicenter studies. These datasets include larger numbers of participants sampled across geographic regions, leading to more diverse cohorts, greater analytic power, and enhanced generalizability. In the United States, the National Alzheimer’s Coordinating Center (NACC) has led a large-scale effort to standardize the collection of common data elements, called the Uniform Data Set (UDS), across more than 35 Alzheimer’s Disease Centers (ADC). The UDS includes a detailed collection of data, such as demographic characterization, clinical scales, self- and partner-reported questionnaires, and importantly for AD/ADRD, a battery of neuropsychological assessments [1].

A core element of clinical phenotyping in AD/ADRD is the accurate and robust measurement of memory. The third version of the UDS (UDS v3.0) neuropsychological battery was released in 2015 [1] and includes eleven cognitive tasks, of which only two primarily assess episodic memory: Craft Story (verbal) and Benson Figure (visual). A notable absence from this battery is a list-learning task, one of the most common [2] and effective memory assessments used by researchers and clinicians. Compared to other episodic memory paradigms, list-learning tasks can provide a more detailed characterization of memory profiles, inform differential diagnosis, and improve prediction of dementia progression [3–6].

Although there are practical benefits to allowing ADCs to vary in list-learning task (e.g., comparison to site-specific historic data), a notable downside is limited cross-site comparability. One example of a project affected by site differences in list-learning procedures is Diverse Vascular Contributions to Cognitive Impairment and Dementia (DVCID). DVCID is a prospective, observational study of vascular brain injury that is focused on Black/African Americans and Latina/o/Hispanic Americans. Its primary goals are to investigate basic mechanisms of small vessel cerebrovascular injury and their relationships with social determinants of health, particularly among individuals of diverse backgrounds [7]. When that study began, the decision was made to allow sites to select from one of six list-learning tasks so they could continue to collect data in a manner consistent with their historic procedures.

The study presented in this manuscript, borne of the desire to develop a harmonized memory metric for the DVCID consortium, sought to leverage best practices in statistical harmonization techniques [8–10] to create a psychometrically robust memory composite that is on the same scale and interpretable regardless of the list-learning task administered. Such a score has utility not only for DVCID, but many studies seeking to include a memory score for cross-AD/ADRD research. This composite will facilitate comparison of data within a study, even if that study changes list-learning tasks over time. In a recent update to the UDS v4.0, studies are required to administer one of two list-learning tasks (both of which are incorporated in the present study) [11]. As a result, sites and studies will differ in which list-learning task they collect. Furthermore, this requirement will introduce differences in list-learning data relative to retrospectively collected UDS v3.0 data. There is therefore a pressing need for a harmonized memory score.

To address this issue, we pooled data from 5943 participants recruited across four consortia and 19 ADCs to create and validate the UDS Memory plus List-Learning (UDS-M+) score, a harmonized memory score that accommodates any of five different list-learning tasks. We then test the performance of this composite in a large independent sample, the DVCID cohort (n=1058). We hypothesize a harmonized memory factor composite score 1) shows adequate model fit and reliability; 2) shows evidence for comparability regardless of contributing list-learning task; 3) demonstrates convergent and discriminate validity with other independent cognitive tasks; and 4) shows evidence of construct validity through associations with disease severity, brain volume, and plasma phosphorylated tau (pTau) levels.

## 2. Methods

### 2.1. Participants

Participants were recruited from 19 ADCs and four AD/ADRD consortia. The score was first developed and tested in a sample of participants (Development cohort; n=5943) recruited through the ARTFL-LEFFTDS Longitudinal Frontotemporal Lobar Degeneration Study (ALLFTD: NCT04363684), Biomarkers for Vascular Contributions to Cognitive Impairment and Dementia (MarkVCID: NCT06284213) and Longitudinal Early-Onset Alzheimer’s Disease Study (LEADS: NCT03507257).

Additional data were also included from the 1Florida, Wisconsin, and University of Southern California ADCs. The UDS-M+ was then tested in a separate cohort of participants enrolled through 16 centers in the DVCID consortium (Validation cohort; DVCID: n=1058). Participants in both cohorts (Table 1) presented with a range of clinical diagnoses and severity levels (including cognitively unimpaired) that were made in consensus conferences using published criteria [12–25]. A subset of participants within the Development cohort provided follow-up data (n=462). Only cross-sectional data was used from the Development Cohort; baseline and follow-up data were included from the Validation cohort.

**Tabel 1:** Sample demographics, clinical diagnoses, and scores for UDS-M+ and components.

|  | <u>Overall</u> | <u>Development</u> | <u>DVCID</u> |
| --- | --- | --- | --- |
| Sample Size | 7001 | 5943 | 1058 |
| Age (years) | 66.57 (11.19) | 65.34 (11.39) | 73.91 (5.85)* |
| Education (years) | 16.01 (2.75) | 16.04 (2.78) | 15.83 (2.59) |
| <u>Race/Ethnicity</u> |  |  |  |
| White non-Hispanic | 2143 (49.0) | 1787 (50.6)* | 356 (42.5) |
| Hispanic or Latino | 499 (11.4) | 313 (8.9) | 186 (22.2)* |
| Black/African American | 753 (17.2) | 464 (13.1) | 289 (34.5)* |
| American Indian or Alaskan Native | 39 (0.9) | 38 (1.1) | 1 (0.1) |
| Pacific Islander | 300 (8.1) | 299 (10.4) | 1 (0.1) |
| Asian | 865 (19.8) | 864 (24.5) | 1 (0.1) |
| Other | 30 (0.7) | 25 (0.7) | 5 (0.6) |
| Female (%) | 3878 (61.0) | 3297 (59.7) | 581 (69.3)* |
| MoCA Total Score | 22.65 (5.71) | 22.13 (5.99) | 24.91 (3.39)* |
| <u>CDR-g (%)</u> |  |  |  |
| 0 | 3049 (50.2) | 2498 (47.7) | 551 (65.8) |
| 0.5 | 2214 (36.4) | 1927 (36.8) | 287 (34.2) |
| 1 | 693 (11.4) | 693 (13.2) | 0 (0.0) |
| 2 | 106 (1.7) | 106 (2.0) | 0 (0.0) |
| 3 | 15 (0.2) | 15 (0.3) | 0 (0.0) |
| CDR-sb | 1.56 (2.41) | 1.75 (2.53)* | 0.38 (0.68) |
| UDS-M+ | 0.04 (0.95) | -0.03 (0.99) | 0.36 (0.73)* |
| <u>Diagnostic Syndrome</u> | <u>Overall</u> | <u>Development</u> | <u>DVCID</u> |
| Cognitively Unimpaired | 1448 (52.0) | 889 (43.6) | 559 (75.1)* |
| Mild Cognitive Impairment | 435 (15.6) | 258 (12.7)* | 177 (23.8) |
| Early-Onset Alzheimer's Dementia | 182 (6.5) | 182 (8.9) | 0 (0.0) |
| Early-Onset Non-Alzheimer's Dementia | 68 (2.4) | 68 (3.3) | 0 (0.0) |
| Alzheimer's Dementia Syndrome | 211 (7.6) | 211 (10.4) | 0 (0.0) |
| Logopenic Variant PPA | 20 (0.7) | 20 (1.0) | 0 (0.0) |
| Dementia with Lewy Bodies | 9 (0.3) | 9 (0.4) | 0 (0.0) |
| Behavioral Variant FTD | 51 (1.8) | 51 (2.5) | 0 (0.0) |
| Amyotrophic Lateral Sclerosis | 10 (0.4) | 10 (0.5) | 0 (0.0) |
| Corticobasal Syndrome | 42 (1.5) | 42 (2.1) | 0 (0.0) |
| Non-Fluent Variant PPA | 25 (0.9) | 25 (1.2) | 0 (0.0) |
| Semantic Variant PPA | 38 (1.4) | 38 (1.9) | 0 (0.0) |
| Progressive Supranuclear Palsy | 27 (1.0) | 27 (1.3) | 0 (0.0) |
| Parkinson's Dementia | 6 (0.2) | 6 (0.3) | 0 (0.0) |
| Vascular Dementia | 4 (0.1) | 4 (0.2) | 0 (0.0) |
| Unspecified Dementia | 8 (0.3) | 0 (0.0) | 8 (1.1) |
| Psychiatric | 12 (0.6) | 12 (1.0) | 0 (0.0) |
| Other | 185 (9.0) | 185 (9.0) | 0 (0.0) |

Presents mean (standard deviation) or count (percent) of demographic variables, diagnoses, and cognitive scores. Data are presented for the full sample, Development cohort, and DVCID.
\* The Development and DVCID cohorts are compared via T-tests or Chi2 tests and the group with significantly larger value is indicated with an asterisk. Comparisons were not completed when the sample size of a cohort was small ( $n < 10$ ).
The "Other" category of clinical diagnoses included unspecified frontotemporal lobar degeneration ( $n=48$ ), multiple system atrophy ( $n=1$ ), unspecified primary progressive aphasia ( $n=14$ ), and those who met no criteria ( $n=94$ ).
Abbreviations: **UDS-M+**: Uniform Data Set Memory plus List-Learning score; **CDR-sb**: Clinical Dementia Rating Scale Sum of Boxes; **CDR-g**: Clinical Dementia Rating Scale Global Score; **MoCA**: Montreal Cognitive Assessment; **HVLT**: Hopkins Verbal Learning Test; **CVLT-II**: California Verbal Learning Test-Version Two; **CVLT-SF**: California Verbal Learning Test Version Two Short Form; **AVLT**: Rey Auditory Verbal Learning Test; **CERAD**: Consortium to Establish a Registry for Alzheimer's Disease; **PPA**: primary progressive aphasia.

#### Inclusion/Exclusion

All participants in both cohorts completed at least one UDS memory test or a list-learning task (see Assessments section). Inclusion criteria required English as the primary language to remove variance associated with language that may impact task performance. An associated study is underway to evaluate the inclusion of tasks completed in other languages. Written informed consent was obtained from all participants or their legal representative. All studies were approved by the Institutional Review Boards of the consortia or ADC in accordance with institutional guidelines and the Helsinki Declaration.

### 2.2. Measures

#### List-learning tasks

Participants were administered one of five list-learning tasks: the California Verbal Learning Test-II, Standard Form (CVLT-II) [26], the California Verbal Learning Test-II, Short Form (CVLT-SF) [26]; Hopkins Verbal Learning Test Revised (HVLT) [27]; Consortium to Establish a Registry for Alzheimer’s Disease Word List Memory Test (CERAD) [28]; and the Rey Auditory Verbal Learning Test (AVLT) [29]. All tests begin with an immediate recall paradigm in which examiners read a list of words to the participant, who are asked to repeat as many words as possible. This immediate recall paradigm is repeated several times. The number of words on the list and the number of immediate recall trials varies by task. The total number of words recalled across all immediate recall trials was used for composite generation (immediate recall). After a pre-specified delay period, which differs by task, participants are asked to freely recall as many words as possible (delayed recall). Participants are then asked to identify correct stimuli in a multiple-choice format (“yes” vs “no”), and a recognition memory discriminability index, *d’*, is generated by integrating the number of correct responses with false positives [26].

#### UDS Memory Measures (Linking Items)

Participants across all list-learning groups completed the UDS Neuropsychological Battery [1], which includes two memory tasks: a story memory task (Craft Story) with verbatim immediate and delayed recall scores; and a visual memory task with a free recall and a recognition trial (correct vs incorrect) of a complex figure (Benson Figure). It also includes the Montreal Cognitive Assessment (MoCA), which provides a brief memory task with a list-learning immediate recall and delayed recall. These three tests provide six memory test items, which are common across all participants.

### 2.3. Other assessments

#### Additional Cognitive Tasks

Participants completed additional UDS measures: Verbal Fluency, Trail Making Test A, Trail Making Test B, Number Span Forward, Number Span Backwards, and the Multilingual Naming Test (MINT) [1, 30]. They also completed a brief cognitive screener (MoCA) [31]. Tasks of executive functioning were summarized into a single executive composite score, the UDS3-EF [32], which uses similar IRT methods to those used to generate the UDS-M+. A subset of participants completed a separate memory task, the Tablet-based Cognitive Assessment Tool (TabCAT) Favorites test [33], an associative memory task involving both visual and verbal stimuli. The outcome was total correct responses across immediate and delayed recall conditions.

#### Clinical Dementia Rating Scale (CDR)

Participants’ functional impairment was rated using the CDR, a clinician-administered semi-structured interview with the participant and a study partner [34]. Clinicians query the following six areas and provide a rating (values: 0, 0.5, 1, 2, 3): memory, judgment and problem solving, community affairs, home and hobbies, orientation, and personal care. The CDR is commonly used in AD/ADRD research and within clinical trials of AD. A weighted algorithm combines scores within all domains into a global score (CDR-g; scores: 0, 0.5, 1, 2, 3) and sum of boxes (CDR-sb; range: 0-18), with higher scores indicating greater impairment.

#### Plasma biomarkers

A subset of Development cohort participants (n=302) provided blood samples at the University of California, San Francisco. Plasma phosphorylated tau-217 (p-tau217) was measured with electrochemiluminescence-based assays on the Meso Scale Discovery platform (MSD, Rockville, MD, USA) using previously published methods [35].

A subset of DVCID participants (n=413) provided blood samples that were processed through DVCID, and the Quanterix Single Molecule Array (SiMoA) assay was used to quantify p-tau181. These methods are fully described elsewhere [7].

#### Imaging

A sample of participants within the Development cohort (MarkVCID; n=385) and Validation cohort (DVCID; n=856) underwent brain MRI with the same acquisition protocol [7]. MRI acquisition and processing is fully described online (https://markvcid.partners.org/markvcid1-protocols-resources) and in prior publications [36]. Imaging correlates of the memory composite were assessed using a priori regions of interests in subsamples with available processed volumetric MRI data. We created a medial temporal lobe ROI comprising bilateral hippocampal, entorhinal, and parahippocampal regions [37–39]. An occipital lobe ROI was created as a control region [40].

To complement the ROI analyses, we leveraged an unbiased whole brain voxel-based morphometry (VBM) approach in a sample of participants at the University of California, San Francisco (n=829) who underwent brain MRI within 90 days of completing the UDS-M+ tasks. These participants were in the Developmental cohort and were distinct from the 385 MarkVCID participants in the imaging analyses described above. Structural MRI data (T1) was acquired with a 3T scanner and processed using Statistical Parametric Mapping in MATLAB (MathWorks, Natick, MA, USA) to conduct a Voxel Based Morphometry (VBM) analysis as previously described [35, 41].

### 2.4. Confirmatory factor analysis with item banking approach for statistical cocalibration

#### Model Building

An item-banking confirmatory factor analysis (CFA) approach to harmonization was conducted as described in detail by Gross et al. 2023 and Vonk et al 2022 [10, 42]. Prior to model creation, continuous variables were recoded into ordinal scores with up to 12 response categories and at least twenty observations in each category to facilitate use of a graded response model approach [43]. The equal interval approach we used to recode variables retains the shape of the distribution of the continuous data. In this study, we divided the sample into five groups based on which list-learning task participants completed: CVLT-II, CVLT-SF, CERAD, HVLT, AVLT. All groups also had the six linking items from the aforementioned UDS Neuropsychological Battery. We then fit a CFA model to one of the subsamples to estimate item thresholds and loadings, including both the linking items and test items specific to that subsample. For the current study, an initial model was fit within the list-learning subsample that had the largest sample size (AVLT). This sample was chosen as its large size may reflect a broader range of ability, although prior research has shown that varying the order of models does not meaningfully affect the quality of factor scores produced through an item-banking CFA [10]. This model resulted in item parameters for the three AVLT variables (immediate recall, delayed recall, and recognition) and all six linking items. A subsequent model was then fit to another subset of data (i.e., a new list-learning task), in which the thresholds and loadings for the linking items—UDS memory tasks in this study—were fixed to their corresponding values from the initial model. Thresholds and loadings for variables from the second list-learning task were freely estimated. This procedure was repeated for all subsamples of the data until parameters were estimated for all list-learning variables. Factor scores from this final model, in which all parameters were fixed, using all available participant data were then estimated to derive a single memory factor score that is on the same scale for each participant regardless of which list-learning task was administered. Factor score creation utilized a Maximum likelihood Robust (MLR) estimator and theta parametrization.

#### Model Fit

The fit of each model was established separately according to factor loadings (>.4 retained [44, 45]), the Tucker Lewis Index (TLI; >0.95 [46]), the Comparative Fit Index (CFI; >0.95 [47]), and the Root Mean Square Error of Approximation (RMSEA; <0.05) [48]. Model building was completed in Mplus [49] via Stata Statistical software [50] using the RUNMPLUS Stata package [51]. All other statistical analyses (e.g., regressions, descriptives) were conducted in R (V4.2.3) [52].

### 2.5. Statistical Analyses

To test for differences between the Development and Validation cohorts on key participant characteristics, linear regressions were fit to compare cohorts on demographic variables, functional impairment, diagnoses, and scores on UDS-M+ component measures (e.g., Craft Story).

#### Comparing scores across list-learning tasks

A primary goal of this project was to create a memory composite that was comparable across participants, regardless of which list-learning task was completed. The average UDS-M+ memory composite in the Development cohort was compared across list-learning samples, after adjusting for differences in sample composition, including CDR-g score, age, education, and sex. Cohen’s *d* effect sizes were calculated for all pairwise comparisons of UDS-M+ memory composites across list-learning groups. Cohen’s *d* magnitudes were categorized as small (≤ 0.20), medium (0.21 to 0.49), and large (≥0.50) [53].

#### Test-Retest reliability

The correlation between baseline and a second visit (retest interval: mean = 12.67 months; SD=1.4; range: 7-18 months) was used to determine test-retest reliability. This analysis was completed in a subset of participants (n=462) who completed the same list-learning task at baseline and follow-up, as well as met at least one of the following conditions: CDR-g=0, MoCA total score >26, or clinical diagnosis of cognitively unimpaired. These sample restrictions were applied to limit the analysis to those least likely to experience disease-related decline while still maximizing input from each list-learning group and study site.

#### Marginal reliability

To examine measurement error of the UDSM+ memory composite across estimated levels of memory ability, marginal reliability was calculated as one minus the square of the standard error of measurement for each participant’s UDS-M+ score [54]. This metric was then plotted against the latent factor to create a test information plot that displays reliability of the latent factor across the latent trait, as measured by the UDS-M+. In general, reliability above .80 is recommended [10]. Test information plots were created for the full sample, then delineated by list-learning group.

### 2.6. Validity analyses

To evaluate evidence for convergent construct validity, separate linear regressions were fit to test the association between the UDS-M+ and each of the following: age, sex, education, and TabCAT Favorites. Each regression adjusted for the CDR-sb, unless CDR-sb was the primary variable of interest. Known-groups validity was tested by fitting linear regressions to examine the effect of CDR-g on the memory composite score. Post-hoc linear regressions investigated the relationship between the UDS-M+ and the CDR memory box score. Discriminant construct validity was evaluated by testing associations with non-memory cognitive tasks [32]. Analyses were first completed in the Development cohort then replicated as possible within the Validation cohort.

Given that memory deficits are an early and common consequence of AD pathology, the association between UDS-M+ score and log (10)-transformed pTau levels were tested via regressions that controlled for age, sex, and education.

The relationship between UDS-M+ and imaging (MTL, occipital) was examined using linear regressions controlling for age, total intracranial volume, education, and CDR-sb. The VBM completed in a subset of the Development cohort included age, total intracranial volume, education, and CDR-sb as covariates. A family wise-error threshold was applied to VBM results (Monte-Carlo; 1000 permutations) to generate an error distribution with a cut off at the Type 1 error threshold (95%). This distribution yields a critical T threshold at a family wise error threshold of p<.05 [55].

## 3. Results

### Sample Characteristics

Participant characteristics are provided in Table 1. Compared to the Development cohort, Validation participants were significantly older and less impaired on the MoCA and CDR. Sample characteristics delineated by list-learning task are presented in Supplemental Table 1.

### Model Building and Fit

In the initial model, all linking item standardized loadings were acceptable (range: 0.60 to 0.93). Linking item loadings were carried forward to all other list-learning models. Fit for all models was adequate or excellent for most fit statistics (CFIs > 0.99: TLIs > 0.99; RMSEA range: 0.03 to 0.09). Fit statistics are summarized in Supplemental Table 2. Unstandardized loadings are presented in Supplemental Table 3 and thresholds for all UDS-M+ variables are presented in Supplemental Table 4.

### Psychometrics and cross-sample comparisons

The UDS-M+ factor score was normally distributed within the full sample and within each list-learning group (Figure 1). After adjusting for differences in sample characteristics (i.e., age, sex, education, and CDR-sb) only the CVLT-II group demonstrated an average Cohen’s *d* value in the medium range (average Cohen’s *d* = -.40). The average Cohen’s *d* estimates were minimal for other list-learning groups (range: −0.05 to −0.24). In participants with longitudinal data (n=462) who were categorized as unimpaired (CDR-g = 0, MoCA > 26, and/or cognitively unimpaired diagnosis), test-retest reliability was adequate (r=0.67; 95% CI: 0.62, 0.72; p<0.001). Marginal reliability was above 0.8 for most participants (93%), only dropping below 0.8 at the highest and lowest ability levels (Figure 2). Supplemental Figure 1 presents marginal reliability delineated by CDR-g groups.

**Figure 1:**
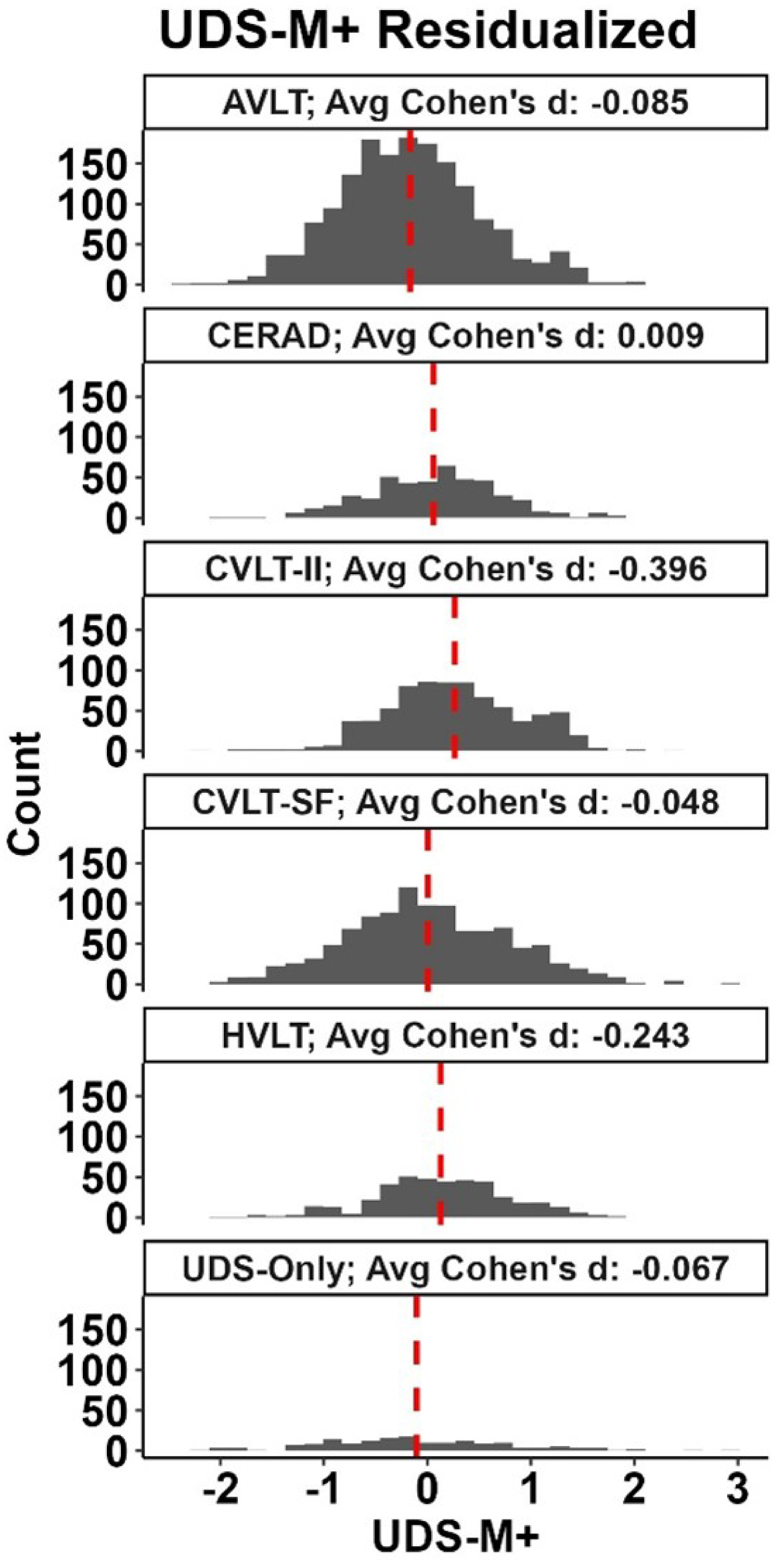
UDS-M+ by list-learning task sample within Development cohort The histograms display the distribution and mean UDS-M+ score (red dashed line) for subsamples delineated by which list-learning task was administered. All subsamples were derived from the Developmental Cohort. T-tests compared UDS-M+ scores (adjusted for age, Clinical Dementia Rating Scale (CDR) sum of boxes, sex, and education) between list-learning subgroups. All combinations of pairwise comparisons were completed. Cohen’s *d*s were calculated for each comparison, and the average Cohen’s *d* is presented above each histogram; for example, −0.085 is the average Cohen’s *d* for pairwise comparisons of the AVLT group and all other list-learning groups. **Abbreviations**: **AVLT**: Rey-Auditory Verbal Learning Test; **CERAD**: Consortium to Establish a Registry for Alzheimer’s Disease; **CVLT-II:** California Verbal Learning Task-II Standard form; **CVLT-SF**: CVLT-II Short form; **HVLT**: Hopkins Verbal Learning Task; **UDS-Only**: Participants who only completed a subset of linking items but were not administered a list-learning task.

**Figure 2:**
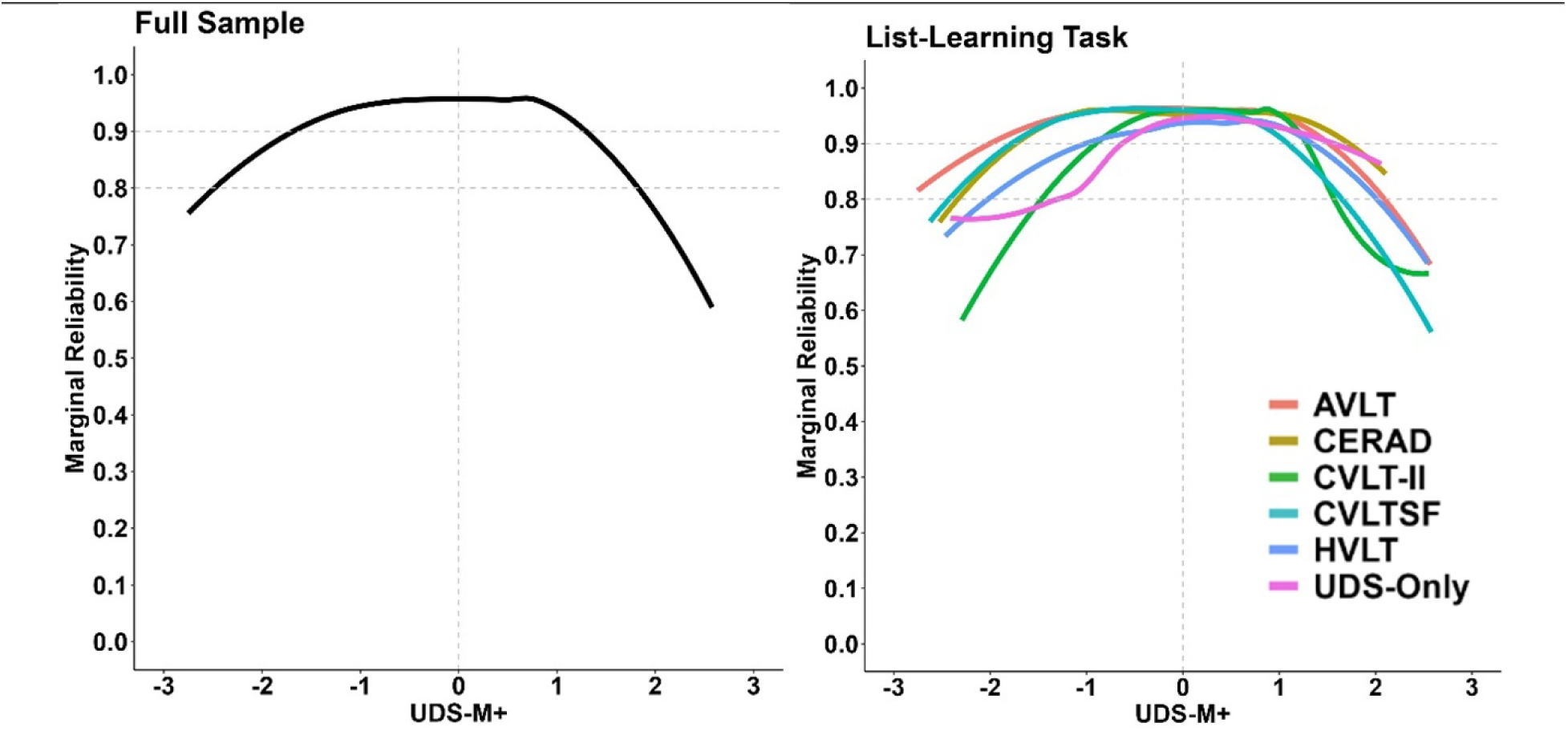
Marginal reliability Presents marginal reliability within a sample that combines the Development cohort. The left graph includes all participants. The right graph separates participants based on which list-learning task they completed. Horizonal dashed lines indicate when the marginal reliability is at 0.8 and 0.9. Abbreviations: AVLT: Rey-Auditory Verbal Learning Test; CERAD: Consortium to Establish a Registry for Alzheimer’s Disease; CVLT-II: California Verbal Learning Task-II Standard form; CVLT-SF: CVLT Short form; HVLT: Hopkins Verbal Learning Task; UDS-Only: Participants who only completed a subset of anchor items.

### Associations with Clinical Measures

Associations between the UDS-M+ and demographics and clinical measures are displayed as standardized regression betas in Figure 3. Within the Development cohort higher UDS-M+ scores were associated as expected with younger age (β=-0.11; 95% CI: −0.09, - 0.13; p<0.001), greater years of education (β=0.12; 95% CI: 0.10, 0.14; p<0.001), and female sex (β=0.17; 95% CI: 0.13, 0.21; p<0.001). Lower scores were associated with greater functional impairment, as measured by the CDR-sb (β=-0.65; 95% CI: −0.67, −0.62; p<0.001) and worse performance on a cognitive screener (MoCA: β=−0.62; 95% CI: -0.60, −0.64; p<0.001). The UDS-M+ was strongly correlated with an independent memory test (TabCAT Favorites: β=0.59; 95% CI: 0.53, 0.66; p<0.001), whereas the magnitude of associations with measures of executive functioning (β range: 0.17, 0.45) and language (β range: 0.21, 0.40) were relatively lower. There was a stepwise decline (Figure 4) in UDS-M+ performance across CDR-g scores (CDR 0 > 0.5 > 1 > 2+; p’s <0.001) and memory box scores (0>1>2+; p<0.001). As shown in Figures 3 and 4, the UDS-M+ associations showed the same general pattern of results in the Development and Validation cohorts. Two associations were notably different between the cohorts. The association between the UDS-M+ and CDR-sb was lower in the Validation cohort than the Development cohort (r = 0.64; 95% CI: 0.62, 0.67; vs r = 0.28 95% CI: 0.24, 0.33). Similarly, the UDS- M+ was less associated with MoCA global scores in the Validation cohort than in the Development Cohort (r = 0.62; 95% CI: 0.60, 0.65 vs r = 0.43; 95% CI: 0.39, 0.46). As noted in Table 1, the cohorts differ in the average and distribution of CDR-sb and MoCA scores, with DVCID representing a less impaired or symptomatic group with restricted variance.

**Figure 3:**
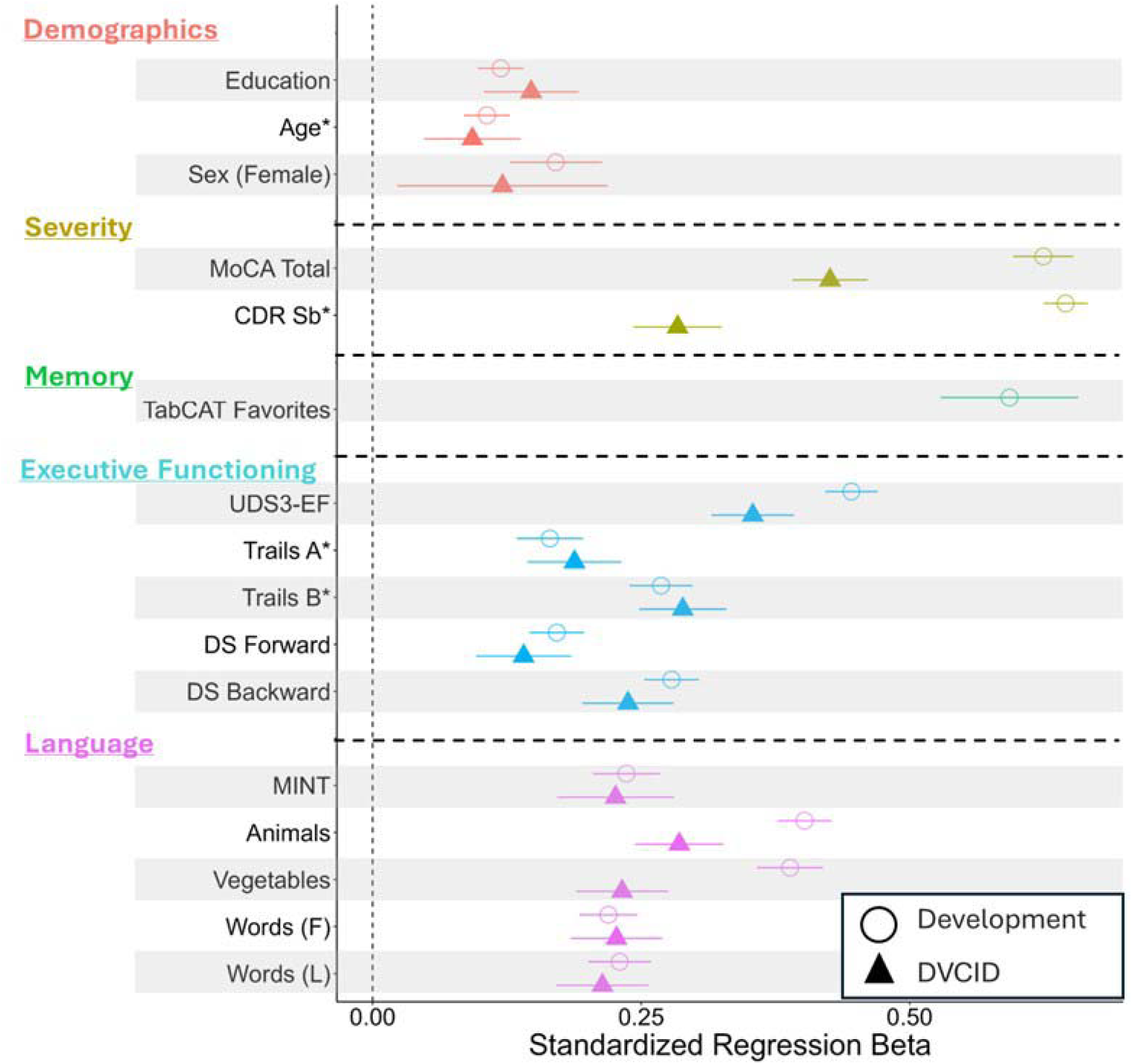
Convergent and discriminant validation Forest plot presenting standardized regression betas in the Development (circle) and DVCID cohorts (triangle), after controlling for CDR Sum of boxes (CDR Sb), with the exception of the CDR Sb row. Confidence intervals (95%) are shown via the horizontal lines. Variables are color coded by domain. * indicates reverse scoring. Abbreviations: MoCA: Montreal Cognitive Assessment total score; TabCAT: Tablet-based Cognitive Assessment Tool; UDS-EF: Uniform Data Set (v3.0) executive function composite score; DS: Digit Span; Words: Lexical Fluency; MINT= Multilingual Naming Task

**Figure 4:**
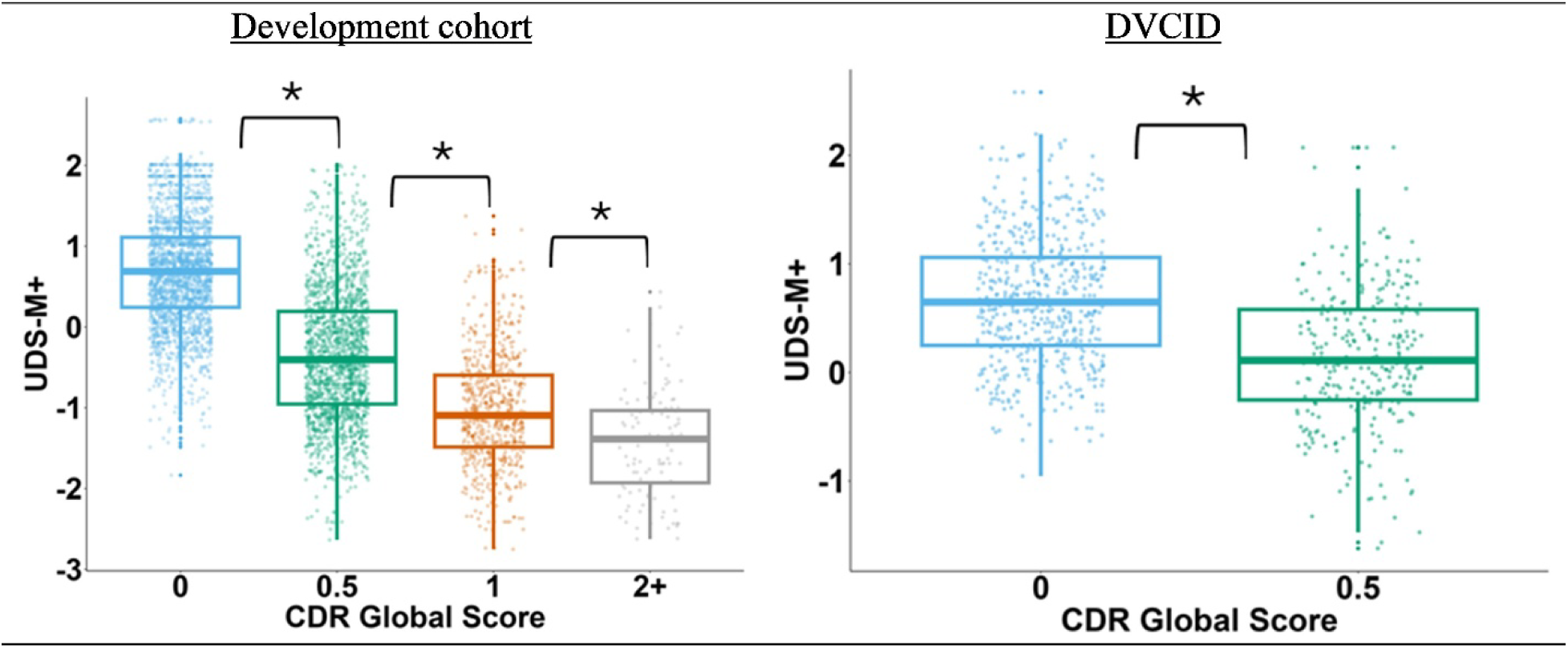
Known Group Validity: UDS-M+ by Clinical Dementia Rating Scale These graphs illustrate the stepwise decline in UDS-M+ scores that was observed with increasing disease stage (p<.001) in two independent cohorts. Abbreviations: UDS-M+: Uniform Data Set Version 3 Memory plus List-Learning score; CDR: Clinical Dementia Rating Scale

### Associations with pTau and brain volume

Worse performance on the UDS-M+ was strongly associated with higher plasma p-tau217 levels in the Development cohort (Figure 5A), after controlling for age, education, sex, and CDR-sb (β=-0.37; 95% CI: −0.46, −0.27; p<0.001). Similarly, in DVCID (Figure 5B), worse performance on the UDS-M+ was associated with higher plasma levels of p-tau181 (β = −0.15; 95% CI: −0.24, −0.05; p=0.002), with the strongest association in the CDR-g = 0.5 group (β = − 0.33; 95% CI: −0.49, −0.17; p<0.001).

**Figure 5:**
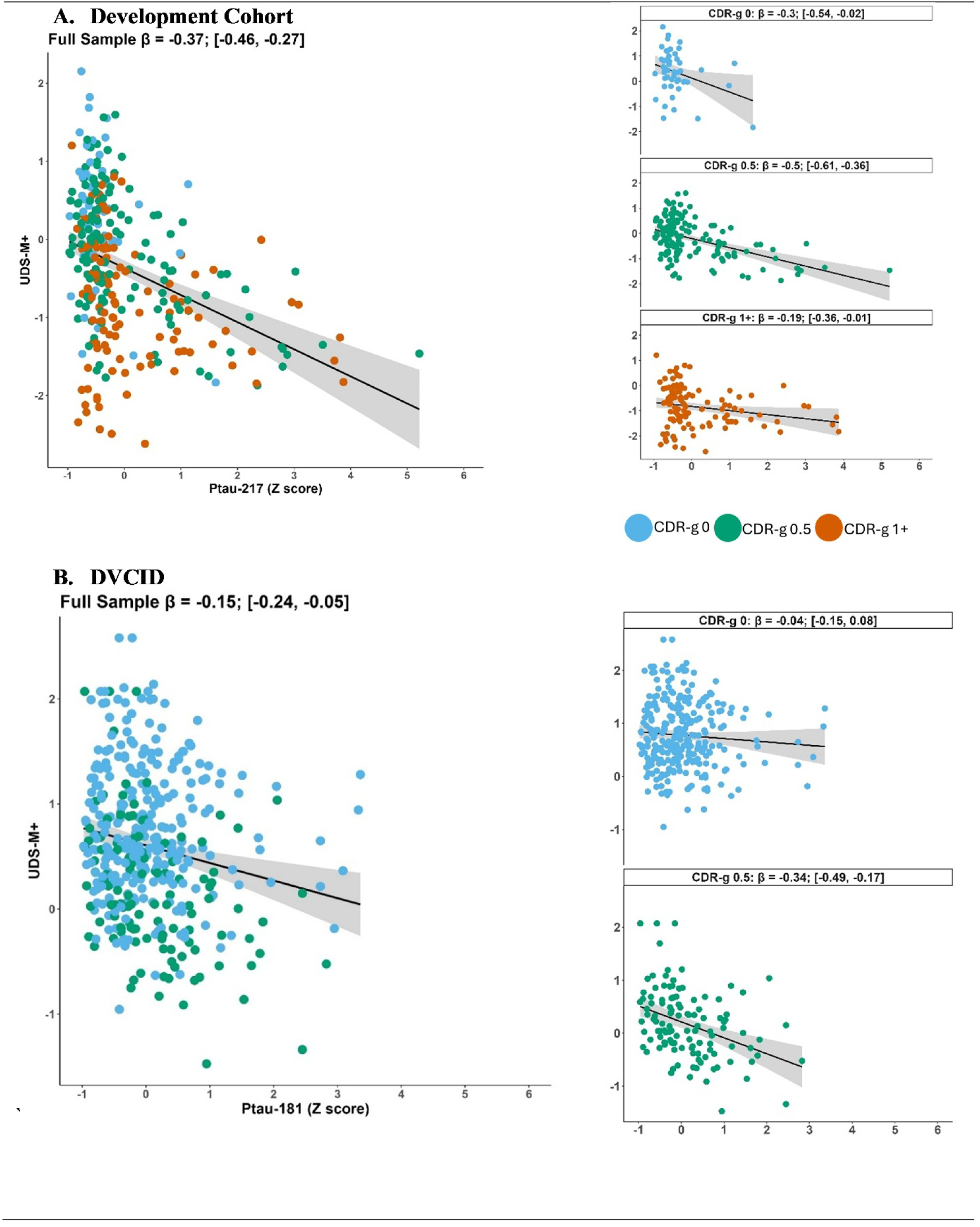
Association between UDS-M+ and plasma phosphorylated tau levels Each graph presents the association between UDS-M+ and plasma phosphorylated tau levels (Development cohort: p-tau217; DVCID: p-tau181), after controlling for age, sex, education, and CDR sum of boxes. Graphs on the left present the results for the full sample within Development cohort (A) and DVCID (B). The stacked graphs on the right display the same results, separated by CDR global scores. p-tau levels were log transformed then Z scored. Abbreviations: UDS-M+: Uniform Data Set Memory plus List-Learning score; CDR: Clinical Dementia Rating Scale

Lower UDS-M+ scores were associated with smaller medial temporal lobe volumes in the Development cohort (β = 0.23 95% CI: 0.13, 0.33; p<.001) and DVCID (β = 0.11 95% CI: 0.04, 0.17; p=0.001). Regarding divergent validity, the UDS-M+ was not significantly associated with the occipital region (β = 0.01 95% CI: −0.05, 0.07; p=.68). In a separate subsample, using a whole-brain VBM approach, worse UDS-M+ scores were associated with smaller brain volumes, with the largest cluster including the medial temporal lobe (Supplemental table 5 and Supplemental figure 2), particularly on the left side of the brain.

## 4. Discussion

We present the development and validation of the UDS-M+, a memory composite score developed using advanced psychometric methods [10, 56] to enable multi-cohort AD/ADRD research. The UDS-M+ was built in a diagnostically heterogeneous sample to support generalizability, and the resulting score showed good model fit with moderate-to-strong factor loadings, and good psychometric properties including a normal distribution and overlapping distributions regardless of the list-learning task administered. Test-retest reliability was adequate and similar to that reported for list-learning tasks over similar time frames [57, 58]. Construct validity was supported by strong associations in the hypothesized directions with demographic factors (e.g., age), an independent memory task, medial temporal lobe volume, and plasma levels of p-tau. The robustness of these findings is bolstered by replication within an independent Validation cohort.

The UDS-M+ was developed using best practices in statistical harmonization, allowing researchers to combine the UDS v3.0 neuropsychological tests and five distinct list-learning tasks into a single composite score. Our results, particularly the minimal residual differences between list-learning task groups (Figure 1) and test information plots (Figure 2), suggest the harmonization process was successful. Importantly, the success of this harmonization is partly demonstrated by the observation that some *differences* in memory composite scores were retained across list-learning tasks and studies. First, the list-learning tasks vary in their length, delay interval, and stimuli. These differences impact the difficulty of the task (i.e., CVLT-II difficulty > CVLT-SF) which in turn affects the relationship between the list-learning task and the UDS-M+. As a result, list-leaning tasks vary somewhat in factor loadings (Supplemental table 3), average UDS-M+ scores (Figure 1), and reliability across levels of memory ability (Figure 2). These variations likely reflect real differences across list-learning tasks and support the use of an IRT-based approach over more simplistic composite approaches (e.g., Z score average) that treat all tasks as equal assessments of the construct. Second, the list-learning tasks were not assigned randomly; for example, nearly all members of a healthy aging study (UCSF: Brain Aging Network for Cognitive Health; BRANCH) completed the CVLT-II Standard Form while all participants of an EOAD study (LEADS) completed the AVLT. A comparison of UDS-M+ scores across these list-learning groups reveals, as expected, a difference in means (Figure 1; CVLT-SF > AVLT) and item reliability across levels of memory (Figure 2).

An important feature of the UDS-M+ is that the resulting memory score is on the same scale regardless of which list-learning task was administered, This feature reflects a significant improvement over the use of raw scores or standardized scores, which are not directly comparable across samples [59–61]. For example, say participant A has a CVLT-II delayed recall of nine words and participant B has a CVLT-SF delayed recall of nine words. These scores do not reflect equal memory abilities despite recalling the same number of words, as there are notable differences in task demands (e.g., word-list length, recall interval, stimuli). An alternative harmonization approach is to convert scores on each list-learning task to Z scores, based on the distribution of the list-learning task within each sample. A complication with Z score comparisons is that the list-learning tasks likely differ in distribution and, like raw scores, do not account for differences in task demands. For example, if participant A (CVLT-II) and participant B (CVLT-SF) both have a Z score of 0 it does not necessarily mean that they have equal memory ability, just that they have average scores within their respective subsamples. The UDS-M+, in contrast, was created with IRT methods that allow for differences in task demands and truly place each participant’s score on the same scale. As a result, if participant A (CVLT-II) had a UDS-M+ score of 0 and participant B (CVLT-SF) had a UDS-M+ score of 0, we can conclude that participant A’s memory is measured to be equal to that of participant B (with a margin of error).

The ability to study memory regardless of list-learning task will become particularly important as the UDS v4.0 requires investigators to administer either the AVLT or CERAD [11]. Thus, many UDS sites who have historically administered a different list-learning task will face a dilemma: they may switch to a new task or administer two list-learning tasks. Switching to a new task disrupts alignment with all retrospectively collected data. Administering two tasks is burdensome for participants and staff and potentially introduces interference effects. This memory composite may offer a solution, as the UDS-M+ can be used to harmonize the AVLT and CERAD with other tasks.

The development of this memory composite adds to several ongoing efforts to advance cognitive research via best practices in statistical harmonization. This item-banking approach has been used to harmonize cognitive instruments administered in six countries through the Health and Retirement Study’s Harmonized Cognitive Assessment Protocol (HRS-HCAP) and has been applied to other longitudinal cohorts [8, 10, 42, 62–64]. AD/ADRD item-banking studies have shown exceptional methodological validation [8, 10, 42, 62, 64], but only a subset have demonstrated convergent validity or included biomarker validation [61, 65]. Our study included both types of validation and provided replication in an independent cohort with racial/ethnic diversity, further highlighting the value of these modern psychometric techniques to AD/ADRD research. Our study also provides a template for other large studies that seek to either combine somewhat disparate cognitive data—for example, various visuospatial tasks—or who wish to alter their data collection. This later point is especially relevant as there is presently a rapid expansion of digital cognitive assessment tools [66–68]; many long-running studies, including the UDS, are modernizing their cognitive batteries and there is a long-overdue push for inclusion of culturally-appropriate cognitive measures [69, 70].

There are limitations to the current version of this score. Marginal reliability was sufficient across all list-learning tasks, though estimates were lower at the high and low extremes of latent memory ability. At least some of the differences in marginal reliability are likely due to sample differences (e.g., disease severity ranges) and task differences (e.g., CVLT-II difficulty > CVLT-SF). As such, it is important for studies that use the UDS-M+ to select list-learning tasks that are appropriate to their sample and, potentially, that vary across participants. For example, the UDS-M+ may better harmonize within a study if impaired participants receive the CVLT-SF and unimpaired participants complete the CVLT-II. Additionally, we primarily present cross-sectional data and have not systematically investigated longitudinal change on the UDS-M+. Ongoing longitudinal analyses will be presented in a separate publication. Although results from the Development cohort support the use of the score across a range of disease severities and diagnoses, and replication in DVCID support its use with participants identifying as Black/African American and Latino/a/Hispanic Americans, additional studies are required to extend the UDS-M+ for use in other testing languages and cultures. Work is underway to develop a version of the score for Spanish speakers. The UDS-M+ was created to aid in comparing memory scores across participants but should not be interpreted as a normatively corrected estimate of memory ability. A score of 0, for example, is relative to this mixed diagnostic group, and should therefore not be interpreted as representing average memory ability at a population level. We also do not provide demographic adjustments, and recommend this step be conducted by investigators in their statistical models based on the relevant confounds in their study. Although the score can accommodate five of the most common list-learning tasks, there are other measures that were not included. The code used to generate the UDS-M+, however, is designed to be iterative, allowing for the addition of other list-learning tasks and memory measures in the future. Interested users with UDS v2.0 data could consider using the crosswalk study to convert Logical Memory to Craft Story scores, although validation is strongly suggested. The code to create the UDS-M+ will be made publicly available at the time of publication.

## Conclusion

We have successfully applied item-banking methods to harmonize data across and within ADCs. The UDS-M+ provides each participant a memory composite score that integrates information across multiple memory measures, regardless of which list-learning task was completed. Without the UDS-M+, ADCs and multisite consortia would be forced to either ignore list-learning data or to combine memory scores in a suboptimal manor [61, 71]. The UDS-M+ may therefore substantially expand and improve the study of memory within ADCs. The successful creation of the UDS-M+ serves as a model for future studies wishing to harmonize internally or who plan to alter their neuropsychological battery during conversion to the UDS v4.0.

## Data Availability

Data from this study are available through the associated consortia. LEADS: https://leads-study.medicine.iu.edu/ MarkVCID: https://markvcid.partners.org/ Diverse VCID: https://diversevcid.ucdavis.edu/ ALLFTD: https://www.allftd.org/

## Acknowledgements

Data were primarily from several consortia encompassing many contributors and sites. We would like to thank all participants and collaborators from the ALLFTD (https://www.allftd.org/), LEADS (https://leads-study.medicine.iu.edu/), MarkVCID (https://markvcid.partners.org/acknowledgements), and DVCID (https://diversevcid.ucdavis.edu/) consortia. We would also like to acknowledge the invaluable contributions of the investigators and support staff at each of the participating sites, including the Administrative, Recruitment and Retention, Statistical and Repository Cores and the DVCID clinical performance sites. We additionally thank those at the 1Flordia, Wisconsin, and USC ADRC.

## Consent Statement

Written informed consent was obtained from all participants or their legal representative. All studies were approved by the Institutional Review Boards of the consortia or ADC in accordance with institutional guidelines and the Helsinki Declaration.

