## Supplement for "Development and validation of a harmonized memory score for multicenter Alzheimer’s disease and related dementia research"

Supplemental Tables and Figures

| **Supplemental table 1: Sample demographics, clinical diagnoses, and scores for UDS-M+ and linking items** | | | | | | | |
| --- | --- | --- | --- | --- | --- | --- | --- |
|  | | **AVLT** | **CVLT-II** | **HVLT** | **CVLTSF** | **CERAD** | **UDS-Only** |
| Sample Size | | 2764 | 1040 | 1063 | 1256 | 686 | 191 |
| Age (years) | | 62.34 (9.22) | 70.33 (9.25) | 72.33 (7.09) | 63.37 (12.53) | 77.01 (7.60) | 60.61 (16.39) |
| Education (years) | | 15.75 (2.60) | 17.01 (2.41) | 15.62 (2.79) | 16.48 (2.82) | 15.39 (2.89) | 15.02 (3.56) |
| Race/Ethnicity | |  |  |  |  |  |  |
| White non-Hispanic | | 1241 (47.1) | 214 (78.1) | 368 (46.6) | 0 (0.0) | 278 (51.5) | 42 (44.7) |
| Hispanic | | 76 (2.9) | 22 (8.0) | 285 (36.1) | 34 (97.1) | 77 (14.3) | 4 (4.3) |
| Black/African American | | 429 (16.3) | 37 (13.5) | 124 (15.7) | 1 (2.9) | 155 (28.7) | 7 (7.4) |
| American Indian | | 35 (1.3) | 0 (0.0) | 2 (0.3) | 0 (0.0) | 2 (0.4) | 0 (0.0) |
| Pacific Islander | | 41 (1.6) | 0 (0.0) | 0 (0.0) | 0 (0.0) | 1 (0.2) | 0 (0.0) |
| Asian | | 802 (30.4) | 1 (0.4) | 2 (0.3) | 0 (0.0) | 19 (3.5) | 41 (43.6) |
| Other | | 13 (0.5) | 0 (0.0) | 9 (1.1) | 0 (0.0) | 8 (1.5) | 0 (0.0) |
| Female (%) | | 1734 (65.6) | 599 (60.0) | 496 (62.8) | 581 (48.7) | 357 (66.1) | 110 (57.9) |
| MoCA Total | | 22.64 (5.69) | 26.10 (2.82) | 24.17 (3.93) | 20.75 (6.40) | 22.48 (5.23) | 18.56 (8.30) |
| CDR-g (%) | |  |  |  |  |  |  |
| 0 | | 1093 (50.3) | 788 (82.6) | 462 (46.4) | 300 (26.5) | 358 (55.9) | 48 (25.9) |
| 0.5 | | 800 (36.8) | 163 (17.1) | 465 (46.7) | 483 (42.7) | 231 (36.1) | 71 (38.4) |
| 1 | | 276 (12.7) | 3 (0.3) | 59 (5.9) | 276 (24.4) | 37 (5.8) | 42 (22.7) |
| 2 | | 3 (0.1) | 0 (0.0) | 8 (0.8) | 65 (5.8) | 13 (2.0) | 17 (9.2) |
| 3 | | 0 (0.0) | 0 (0.0) | 1 (0.1) | 6 (0.5) | 1 (0.2) | 7 (3.8) |
| CDR-sb | | 1.49 (2.00) | 0.21 (0.64) | 1.09 (1.80) | 3.11 (3.03) | 1.17 (2.25) | 4.03 (4.39) |
| UDS-M+ | | 0.04 (0.98) | 0.71 (0.71) | 0.29 (0.81) | -0.17 (1.00) | 0.08 (0.89) | -0.57 (1.07) |
| **Diagnostic Syndrome** | | | | | | | |
| CU | 227 (43.3) | | 694 (83.7) | 257 (79.6) | 207 (21.0) | 63 (85.1) | 0 (0.0) |
| MCI | 53 (10.1) | | 100 (12.1) | 61 (18.9) | 205 (20.7) | 10 (13.5) | 5 (11.6) |
| EOAD | 172 (32.8) | | 0 (0.0) | 0 (0.0) | 0 (0.0) | 0 (0.0) | 10 (23.3) |
| EOAD Non-AD | 68 (13.0) | | 0 (0.0) | 0 (0.0) | 0 (0.0) | 0 (0.0) | 0 (0.0) |
| AD | 0 (0.0) | | 0 (0.0) | 0 (0.0) | 205 (20.7) | 0 (0.0) | 6 (14.0) |
| lvPPA | 0 (0.0) | | 0 (0.0) | 0 (0.0) | 19 (1.9) | 0 (0.0) | 1 (2.3) |
| DLB | 0 (0.0) | | 0 (0.0) | 0 (0.0) | 9 (0.9) | 0 (0.0) | 0 (0.0) |
| bvFTD | 0 (0.0) | | 0 (0.0) | 0 (0.0) | 48 (4.9) | 0 (0.0) | 3 (7.0) |
| ALS | 0 (0.0) | | 0 (0.0) | 0 (0.0) | 10 (1.0) | 0 (0.0) | 0 (0.0) |
| CBS | 0 (0.0) | | 0 (0.0) | 0 (0.0) | 39 (3.9) | 0 (0.0) | 3 (7.0) |
| nvPPA | 0 (0.0) | | 0 (0.0) | 0 (0.0) | 24 (2.4) | 0 (0.0) | 1 (2.3) |
| svPPA | 0 (0.0) | | 0 (0.0) | 0 (0.0) | 37 (3.7) | 0 (0.0) | 1 (2.3) |
| PSP | 0 (0.0) | | 0 (0.0) | 0 (0.0) | 27 (2.7) | 0 (0.0) | 0 (0.0) |
| PDD | 0 (0.0) | | 1 (0.1) | 0 (0.0) | 5 (0.5) | 0 (0.0) | 0 (0.0) |
| VD | 0 (0.0) | | 0 (0.0) | 0 (0.0) | 3 (0.3) | 0 (0.0) | 1 (2.3) |
| Unspecified | 2 (0.4) | | 0 (0.0) | 5 (1.5) | 0 (0.0) | 1 (1.4) | 0 (0.0) |
| Psychiatric | 0 (0.0) | | 0 (0.0) | 0 (0.0) | 12 (1.2) | 0 (0.0) | 1 (2.3) |
| Other | 2 (0.4) | | 34 (4.1) | 0 (0.0) | 138 (14.0) | 0 (0.0) | 11 (25.6) |
| **UDS-M+ Linking Items (mean (SD))** | | | | | | | |
| MoCA Immediate Recall | 8.64 (2.08) | | 9.52 (1.10) | 8.74 (1.98) | 8.22 (2.48) | 8.58 (1.76) | 6.66 (3.28) |
| MoCA Delayed Recall | 2.25 (1.86) | | 3.12 (1.56) | 2.83 (1.71) | 1.64 (1.78) | 2.42 (1.79) | 1.66 (1.91) |
| Craft Immediate Recall | 16.94 (8.45) | | 20.57 (7.24) | 18.95 (7.44) | 14.56 (8.68) | 17.97 (7.58) | 12.71 (8.42) |
| Craft Delayed Recall | 13.93 (8.78) | | 17.83 (7.48) | 15.66 (7.78) | 11.80 (8.89) | 14.58 (8.31) | 9.67 (8.57) |
| Benson Delay Recall | 9.05 (4.77) | | 11.54 (3.14) | 9.41 (3.65) | 8.03 (4.91) | 8.30 (4.40) | 7.10 (5.34) |
| Benson Recognition | 0.78 (0.41) | | 0.90 (0.29) | 0.87 (0.34) | 0.74 (0.44) | 0.75 (0.43) | 0.69 (0.47) |
| Presents mean (standard deviation) or count (percent) of demographic variables, diagnoses, and cognitive scores. Data are combined from the Developmental and DVCID cohorts. The “Other” category of clinical diagnoses included unspecified frontotemporal lobar degeneration (n=48), multiple system atrophy (n-1), unspecified primary progressive aphasia (n=14), and those who met no criteria (n=94).  Abbreviations: **CDR-sb**: Clinical Dementia Rating Scale Sum of Boxes; **CDR-g**: Clinical Dementia Rating Scale Global Score; **MoCA**: Montreal Cognitive Assessment; **HVLT**: Hopkins Verbal Learning Test; **CVLT-II**: California Verbal Learning Test-Version Two; **CVLTSF**: California Verbal Learning Test Version Two Short Form; **AVLT**: Rey Auditory Verbal Learning Test; **CERAD**: Consortium to Establish a Registry for Alzheimer's Disease; **CU**: cognitively unimpaired; **MCI**: Mild Cognitive Impairment; **AD**: Alzheimer’s dementia; logopenic primary progressive aphasia; **DLB**: dementia with Lewy Bodies; **bvFTD**: behavioral variant Frontotemporal Dementia; **ALS**: Amyotrophic lateral sclerosis; **CBS**: Corticobasal Syndrome; nvPPA: nonfluent PPA; svPPA: semantic variant PPA; **PSP**: Progressive Supranuclear Palsy; **PDD**: Parkinson’s Dementia; **VD**: Vascular Dementia. | | | | | | | |

| **Supplemental Table 2: Model fit statistics** | | | |
| --- | --- | --- | --- |
|  | RMSEA | CFI | TLI |
| AVLT [Initial Model] | 0.09 | 0.995 | 0.992 |
| CVLT-II | 0.033 | 0.997 | 0.996 |
| HVLT | 0.048 | 0.996 | 0.994 |
| CVLT-SF | 0.084 | 0.993 | 0.990 |
| CERAD | 0.057 | 0.997 | 0.995 |
| Presents fit statistics for each of the five list-learning models. Each fit statistic is interpreted as adequate based on prior guidelines; RMSEA; <.05 [42]; CFI; >0.95 [41]; TLI; >0.95 [40]. **Abbreviations**: **AVLT**: Rey-Auditory Verbal Learning Test; **CERAD**: Consortium to Establish a Registry for Alzheimer's Disease; **CVLT-II:** California Verbal Learning Task-II Standard form; **CVLTSF**: CVLT-II Short form; **HVLT**: Hopkins Verbal Learning Task; **UDS-Only**: Participants who only completed a subset of anchor items but were not administered a list-learning task; **RMSEA**: Root mean square error of approximation; **CFI**: Comparative fit index ; **TLI**: Tucker-Lewis index . | | | |

| **Supplemental Table 3: Unstandardized loadings** | | |
| --- | --- | --- |
| Measure | UDS-M+ Loading | Task-Specific Loading |
| MoCA Immediate | 0.99 | -- |
| MoCA Delayed | 1.67 | -- |
| Benson Recall | 1.42 | -- |
| Benson Recognition | 0.76 | -- |
| Craft Immediate | 2.65 | 1.963 |
| Craft Delayed | 3.18 | 1.963 |
| AVLT Immediate | 2.48 | -0.463 |
| AVLT Delayed | 2.88 | -0.463 |
| AVLT Recognition | 1.51 | -0.463 |
| CVLT-II Immediate | 3.00 | 0.767 |
| CVLT-II Delayed | 3.55 | 0.767 |
| CVLT-II Recognition | 2.83 | 0.767 |
| HVLT Immediate | 2.28 | -0.685 |
| HVLT Delayed | 1.98 | -0.685 |
| CVLT-SF Immediate | 1.58 | -0.244 |
| CVLT-SF Delayed | 2.75 | -0.244 |
| CVLT-SF Recognition | 1.92 | -0.244 |
| CERAD Immediate | 2.29 | 1.000 |
| CERAD Delayed | 3.32 | 0.923 |
| CERAD Recognition | 1.91 | 0. 923 |
| Presents the unstandardized loadings between observed variables and latent factors. The “UDS-M+ Loading” column displays the loading between that variable and the overall memory factor composite score. The “Task-Specific Loading” displays the loading between each variable and a latent factor meant to account for test-specific variance between similar items. Test-Specific factors were included for the Craft Story and each list-learning task. Test-Specific factors were not included for the MoCA or the Benson due to model fitting errors.  **Abbreviations**: **AVLT**: Rey-Auditory Verbal Learning Test; **CERAD**: Consortium to Establish a Registry for Alzheimer's Disease; **CVLT-II:** California Verbal Learning Task-II Standard form; **CVLTSF**: CVLT-II Short form; **HVLT**: Hopkins Verbal Learning Task; | | |

| **Supplemental Table 4: Thresholds for all UDS-M+ items** | | | | | | |
| --- | --- | --- | --- | --- | --- | --- |
| Binned Score | MoCA Immediate | MoCA Delayed | Craft Immediate | Craft  Delay | Benson  Delay | Benson Recognition |
| 1 | -1.250 | -0.837 | -5.293 | -3.424 | -1.941 | -0.905 |
| 2 | -0.073 | -0.364 | -3.412 | -2.246 | -1.502 |  |
| 3 |  | 0.269 | -2.212 | -1.312 | -1.326 |  |
| 4 |  | 1.040 | -1.217 | -0.311 | -0.838 |  |
| 5 |  |  | 0.000 | 1.229 | -0.595 |  |
| 6 |  |  | 1.026 | 2.396 | -0.077 |  |
| 7 |  |  | 2.063 | 3.676 | 0.195 |  |
| 8 |  |  | 3.558 | 4.984 | 1.06 |  |
| 9 |  |  | 4.751 | 6.674 | 1.543 |  |
| Binned Score | AVLT Immediate | AVLT Delay | AVLT Recognition | CVLT-II Immediate | CVLT-II Delayed | CVLT-II Recognition |
| 1 | -6.032 | -2.267 | -3.777 | -2.067 | -2.186 | -0.312 |
| 2 | -4.612 | -1.654 | -2.942 | -1.391 | -1.056 | 0.579 |
| 3 | -3.368 | -0.756 | -2.409 | -0.499 | 0.198 | 1.389 |
| 4 | -2.200 | -0.362 | -1.776 | 0.417 | 0.679 | 2.275 |
| 5 | -1.130 | 0.103 | -1.311 | 1.243 | 2.14 |  |
| 6 | 0.285 | 1.064 | -0.962 | 1.999 | 3.451 |  |
| 7 | 0.604 | 1.696 | -0.526 | 2.937 |  |  |
| 8 | 1.788 | 2.360 | -0.056 | 3.826 |  |  |
| 9 | 3 .157 | 3.976 |  |  |  |  |
| Binned Score | CERAD Immediate | CERAD Delay | CERAD Recognition | CVLT-SF Immediate | CVLT-SF Delayed | CVLT-SF Recognition |
| 1 | -3.394 | -2.365 | -4.543 | -3.602 | -2.806 | -4.62 |
| 2 | -2.440 | -1.917 | -3.394 | -2.854 | -2.272 | -3.727 |
| 3 | -1.812 | -1.255 | -2.275 | -2.092 | -1.841 | -3.221 |
| 4 | -1.445 | -0.750 |  | -1.594 | -1.319 | -2.667 |
| 5 | -0.655 | 0.093 |  | -0.885 | -0.825 | -2.183 |
| 6 | 0.186 | 0.915 |  | -0.171 | -0.266 | -1.582 |
| 7 | 0.651 | 2.090 |  | 0.336 | 0.400 | -1.075 |
| 8 | 1.466 | 3.105 |  | 1.226 | 1.398 | -0.218 |
| 9 | 2.484 |  |  | 2.259 |  |  |
|  |  | Binned Score | HVLT Immediate | HVLT Delay |  |  |
|  |  | 1 | -3.528 | -0.360 |  |  |
|  |  | 2 | -2.272 | -0.167 |  |  |
|  |  | 3 | -0.995 | 0.173 |  |  |
|  |  | 4 | 0.068 | 0.427 |  |  |
|  |  | 5 | 1.407 | 0.723 |  |  |
|  |  | 6 |  | 1.575 |  |  |
|  |  | 7 |  | 2.083 |  |  |
|  |  | 8 |  |  |  |  |
|  |  | 9 |  |  |  |  |
| Presents the thresholds produced from the item response theory structural equation models. The “Binned Score” column indicates the recoded, ordinal value for each task. Each column indicates a specific task. **Abbreviations**: **AVLT**: Rey-Auditory Verbal Learning Test; **CERAD**: Consortium to Establish a Registry for Alzheimer's Disease; **CVLT-II:** California Verbal Learning Task-II Standard form; **CVLTSF**: CVLT-II Short form; **HVLT**: Hopkins Verbal Learning Task. | | | | | | |
| **Supplemental Figure 1: Marginal Reliability for CDR-g groups.** | | | | | | |
| 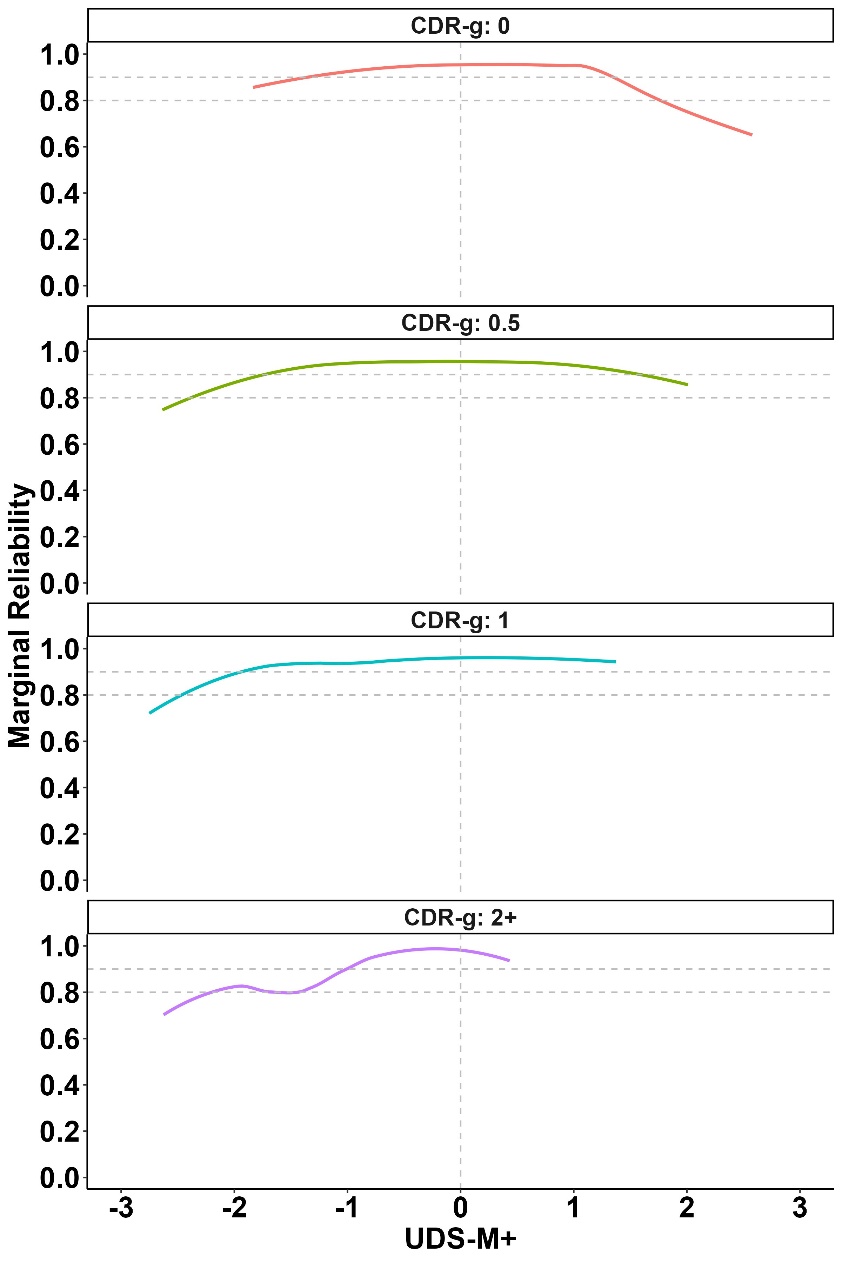 | | | | | | |
| Presnts marginal reliabilty delinated by Clincal Dementia Rating scale (CDR) global scores. Dotted horizontal gray lines highlight marginal reliability values above 0.80 and 0.90. A vertical gray line designates average UDS-M+ scores within the overall sample, indicating average latent ability (memory). | | | | | | |

| **Supplemental Table 5: Voxel-based Morphometry results** | | | | | |
| --- | --- | --- | --- | --- | --- |
| Region | Volume (mm^3)^ | X | Y | Z | Max T |
| L. Middle temporal gyrus | 133772 | -62 | -40 | -15 | 13.47 |
| R. Middle frontal gyrus | 97 | 28 | 3 | 58 | 5.48 |
| R. Frontal pole | 62 | 14 | 68 | 8 | 5.04 |
| R. Postcentral gyrus | 54 | 30 | -38 | 63 | 4.94 |
| R. Middle frontal gyrus | 32 | 30 | 36 | 42 | 4.99 |
| R. Inferior frontal gyrus | 15 | 45 | 48 | 9 | 4.67 |
| R. Anterior orbital gyrus | 14 | 24 | 32 | -20 | 4.81 |
| L. Superior frontal gyrus | 10 | -24 | -10 | 64 | 4.70 |
| Presents regions with T values > 4.36, which is the critical T based on the permutation method, and a volume >= 10 mm^3^. The x, y, and z columns provide MNI coordinates of the most significant voxel within that region. “L” indicates left side; “R” indicates right side. | | | | | |

**Supplemental figure 2: Voxel wise associations of the UDS-M+ with brain volume**

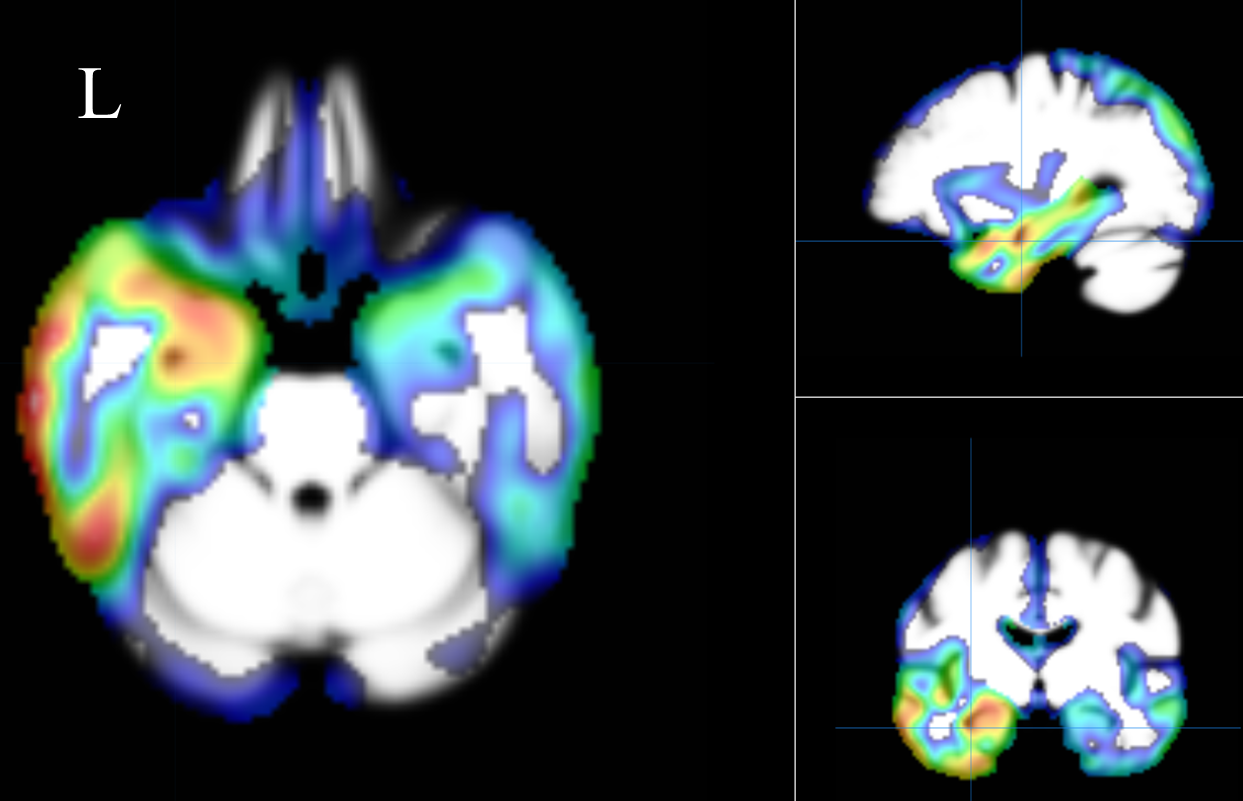

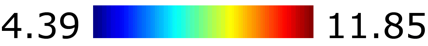

Presents voxel-based morphometry brain correlates of the UDS-M+ in a subsample (n=829) of Development cohort participants. The “L” denotes the left-right orientation of the images. The color bar indicates the range of critical T values that remained statistically significant after family-wise error correction (p<.05). Crosshairs are centered on the left hippocampus. The UDS-M+ was strongly associated with several brain regions, particularly within the left hemisphere, including the amygdala, temporal gyrus (medial > inferior > superior), hippocampus, fusiform gyrus, insula, and temporal pole.
